# Impact of a Pharmacy Care Management Service on Cardiometabolic Medication Adherence and Resource Use for Medicare Advantage Beneficiaries

**DOI:** 10.1101/2025.02.18.25322491

**Authors:** Shawn Hallinan, Loren Lidsky, Josh Benner, Stephen Jones, Elise Smith, Chronis Manolis, Chester B Good, Niteesh K. Choudhry

## Abstract

**BACKGROUND:** Prescribing rates and adherence to evidence-based cardiometabolic medications remain suboptimal. Many strategies to improve prescribing and adherence have been developed; pharmacy care management (PCM) programs are among the most consistently effective of these. The clinical and economic benefits of ongoing PCM programs for patients with cardiometabolic conditions are incompletely understood.

**OBJECTIVES:** To evaluate the effect of a PCM program in individuals with cardiometabolic conditions, specifically, whether the program improved adherence to cardiometabolic medications and reduced all-cause and cardiometabolic-specific healthcare use, and whether those non-adherent at baseline would benefit most.

**METHODS:** We conducted a retrospective cohort study using adjudicated administrative claims data from a large regional Medicare Advantage Prescription Drug (MAPD) health plan. The PCM program was offered to MAPD beneficiaries who filled ≥ 8 chronic medications in the 180 days prior to eligibility screening. We restricted the cohort to individuals who filled ≥ 2 prescriptions for a cardiometabolic condition, filled at least one prescription after PCM enrollment, and were continuously eligible for health plan benefits for at least 12 months before and after enrollment. Potential controls were patients who met the same criteria but did not participate in the PCM program and filled prescriptions at non-PCM pharmacies. Control patients were matched to PCM patients in a 5:1 ratio using direct and propensity score matching. The primary outcome was all-cause hospitalization during the 12-month follow-up period. Secondary outcomes included cardiometabolic medication adherence and disease-specific hospitalizations. In pre-planned stratified analyses, we further evaluated the program effects on rates of disease-specific hospitalization in patients who were non-adherent versus adherent at baseline.

**RESULTS:** The final cohort consisted of 632 PCM patients and 3,160 well-matched controls. Compared to controls, PCM program participants had significantly higher rates of adherence to all classes of cardiometabolic medications, ranging from 4.7% (P = 0.028) for anticoagulants to 17.0% (P < 0.001) for beta-blockers. PCM program participation had 15% fewer all-cause hospitalizations per 1,000 patient months (P = 0.037) than matched controls. PCM patients experienced significantly fewer cardiometabolic-specific admissions (-33.7%; P = 0.025) and non-significant reductions in non-cardiometabolic admissions (-8.4%; P = 0.201). PCM patients who were non-adherent at baseline had a significant 39.1% reduction in cardiometabolic hospitalizations (P = 0.003) from baseline, while adherent patients had a non-significant 24.7% reduction (P = 0.286).

**CONCLUSION:** Compared with closely matched controls, PCM patients with pre-existing cardiometabolic disease had significantly higher rates of medication adherence and significantly lower hospitalization rates during the 12-month follow-up period. This effect was greatest in patients who were non-adherent at baseline. Our results provide insights into how PCM programs achieve their benefits and underscore the value of targeting the PCM program to high-risk individuals.

---

Numerous medications for the prevention of cardiometabolic morbidity and mortality and the management of risk factors have been developed, rigorously tested, approved for use, and have become the standard of care for patients with cardiometabolic disease.^1^ Unfortunately, prescribing rates for these evidence-based drugs remain suboptimal; among patients initiating therapy, many do not fill their initial prescriptions, and among those who do, long-term adherence remains low.^2–4^

Many strategies to improve cardiometabolic medication use have been developed and tested. Among the most consistently effective are pharmacist-led multicomponent interventions that include medication reviews, patient counselling, recommendations for medication changes to physicians, and reminder devices or packaging solutions.^5–10^ The majority of data evaluating this approach come from studies that developed interventions specifically for research purposes rather than from real-world programs delivering integrated, comprehensive clinical pharmacy services to patients who need them.^11^

We previously evaluated an existing pharmacy care management (PCM) program and found it improved medication adherence, reduced hospitalizations, and lowered overall spending among medically complex and vulnerable Medicare beneficiaries with a broad range of health conditions.^12^ Given the particular and persistent burden of cardiometabolic disease in the US,^13,14^ the current evaluation specifically assessed the program’s effect for individuals with these conditions. In addition, to help understand the mechanism by which PCM programs work, we evaluated whether the overall benefit of the program was: (1) attributable to improvements in cardiometabolic outcomes or to more general efforts to improve evidence-based prescription drug use and (2) greater among those anticipated to benefit most, specifically those who were non-adherent to their prescribed cardiometabolic medications prior to joining the program.^12^

## METHODS

### Setting and design

This retrospective cohort study evaluated the impact of a PCM program on medication adherence and health services utilization for individuals with cardiometabolic disease enrolled in the University of Pittsburgh Medical Center (UPMC) Medicare Advantage program. This study was a program evaluation by UPMC Health Plan and thus deemed exempt by the UPMC Institutional Review Board.

### Subject eligibility and study cohorts

The PCM program was offered to beneficiaries of UPMC Medicare Advantage plans who filled eight or more medications intended for chronic use in the 180 days prior to being screened for eligibility. Eligibility rules for the PCM program excluded individuals enrolled in a state-sponsored pharmaceutical assistance program or using a pharmacy known to provide services like the PCM program. The current analysis was restricted to individuals who had filled at least 2 prescriptions for a cardiometabolic condition (**Table e1**) in the 24 to 13-month period before being screened for eligibility and who had maintained insurance eligibility for 12 months before and 12 months after being screened. We excluded individuals in hospice at the time of potential PCM program eligibility.

Patients were screened for eligibility and offered the PCM program by telephone on a rolling basis starting in December 2019; the last date of enrollment for patients included in this evaluation was November 2021. The PCM cohort consisted of individuals who agreed to enroll in the PCM program, filled at least one prescription after enrollment, and remained enrolled for at least 12 months. Potential controls were beneficiaries eligible for the PCM program who could not be reached or were reached and declined to enroll and subsequently filled a medication at a non-PCM pharmacy. To reduce the potential for selection bias, we excluded potential controls who declined participation and asked not to be contacted for potential enrollment again. The follow-up period began on the index date, which was the date of the first prescription fill after enrollment (PCM patients) or the first prescription fill after outreach was attempted (controls).

### PCM intervention

After agreeing to enroll, patients participated in an onboarding appointment by telephone to obtain medication history, reconcile current prescriptions, and synchronize medication refills. Clinical pharmacy services and medication dispensings were provided by a single pharmacy, Mosaic Pharmacy Service (Sterling, Virginia; a subsidiary of RxAnte). Medications were delivered to patients’ homes in specialized dispensing packaging in 30-day cycles to minimize waste compared with extended-day supplies, given that medications for this population change frequently. Medications were sorted into pouches corresponding to the day and time of each dose. A large label with photographs and instructions for each medication was affixed to the box containing the monthly supply of pouches.

Subsequently, all patients were contacted monthly by a pharmacist or certified pharmacy technician to assess changes to health status and their medication regimen. On an as-needed basis, clinical pharmacists consulted with the patients’ physicians, provided education, and made referrals to health care providers and social support or case management programs provided by the Medicare Advantage plan.

### Baseline covariates

We measured baseline characteristics in the 12-month period prior to the index date for all PCM and potential control patients. These included age, gender, index date, receipt of the Medicare low-income subsidy, number of chronic medications, Charlson Comorbidity Index score, health services use (i.e., emergency room visits and hospitalizations), total inpatient days, baseline pharmacy spending, baseline medical spending, and the proportion of patients who completed a comprehensive medical review in the past 12 months. We calculated patients’ predicted cost savings from improved medication adherence using the Value of Future Adherence (VFA) score. VFA is a commercially available algorithm (developed by RxAnte, Inc.) that predicts potential 12-month cost savings from improving medication adherence.^16–18^ The VFA score in the study population ranged from $0 to $7,000. We defined *high VFA* as a score of ≥ $3,000, which represented the top quartile of patients.

We also calculated baseline adherence for each class of cardiometabolic medications, specifically renin-angiotensin-system-acting agents, beta blockers, calcium channel blockers, cholesterol lowering medications, oral anticoagulants, antiplatelets, and oral hypoglycemics, by calculating the proportion of days covered during the 12-month baseline period using methods consistent with the Centers for Medicare & Medicaid Services Medicare Star Ratings measure specifications.^15^ Accordingly, to be eligible for the adherence evaluation, patients had to have filled two or more prescriptions in the drug class at any point during the 12-month period prior to the baseline period, and patients were considered adherent if their PDC for a given class was ≥ 80% during the baseline period.

### Creation of matched cohorts

We matched PCM patients with potential controls on variables associated with PCM enrollment, healthcare utilization, and medication adherence. First, we directly matched PCM to potential controls on age, total pre-index hospitalizations, pre-index total medical costs and total pharmacy costs, VFA score, and completion of a comprehensive medical review. After this, we propensity matched exactly 5 potential controls to each PCM patient based on 8 additional variables: days since index date, sex, low-income subsidy status, Charlson Comorbidity Index score, 90-day supply of and total unique maintenance medications, emergency room visits, and total inpatient days. We used a greedy matching approach, with calipers of width equal to 0.2 of the standard deviation of the logit of the propensity score.^19–21^ The effectiveness of the match was assessed with standardized mean differences (SMD), with an SMD of <10% considered well-matched,^22,23^ and post-match c-statistics, where values approaching 0.5 suggest measured characteristics of a PCM patient and a control patient are indistinguishable.^24^

### Outcomes

Our primary outcome was all-cause hospitalizations during the 12-month follow-up period as assessed using adjudicated claims data. We also evaluated all-cause emergency room and urgent care visits and medication adherence during the follow-up period. Medication adherence was calculated separately for each therapy class. Similar to the approach taken for baseline adherence, follow-up adherence was calculated among patients who filled two or more prescriptions in the drug class at any point during the 12-month period prior to the baseline period. Patients were considered adherent if their PDC for a given class was ≥ 80% during the follow-up period.

We used primary diagnosis codes for hospitalizations to stratify admissions into cardiometabolic (ICD-10 I00-I99, E00-E89) and non-cardiometabolic categories.

### Statistical analyses

Descriptive statistics were used to characterize the study cohort before and after propensity-score matching. To adjust for any residual baseline imbalances between the PCM and control groups after matching, a difference-in-difference approach was used to calculate a relative difference in all-cause hospitalization. This was defined as the ratio of the absolute difference between follow-up and baseline PCM utilization, adjusted for 12-month pre-post changes in control utilization. We used bootstrapping (10,000 iterations with replacement) to test the statistical significance of the difference between PCM and control based on a one-sided alpha level of 0.05. An identical approach was used for other measures of healthcare utilization. Absolute differences in adherence between PCM and control patients were estimated using a Poisson generalized linear model with log link and robust standard errors that adjusted for pre-period adherence.

To explore how PCM may impact outcomes, we stratified hospitalizations into cardiometabolic and non-cardiometabolic specific causes and re-ran our difference-in-difference models. We then further stratified cardiometabolic specific hospitalizations based on whether patients were non-adherent or adherent to their prescribed cardiometabolic medications prior to joining the program.

All analyses were conducted by using SAS version 9.4 (SAS Institute, Inc., Cary, North Carolina).

## RESULTS

The study cohort consisted of 657 PCM patients and 14,014 potential controls (**Figure 1**), whose baseline characteristics prior to matching are shown in **Table e2**. After two-stage matching, PCM and control patients were well-matched with respect to baseline characteristics, with all SMDs less than 10% and a post-match c-statistic of 0.53 (**Table 1**). Patients had a mean age of 69 (10.1) years, 39% were male, 41% received a low-income subsidy, and the average Charlson Comorbidity Index score was 3.6. Patients were taking an average of 11 maintenance medications, and 98% had filled at least one 90-day supply in the baseline period.

**Figure 1.**
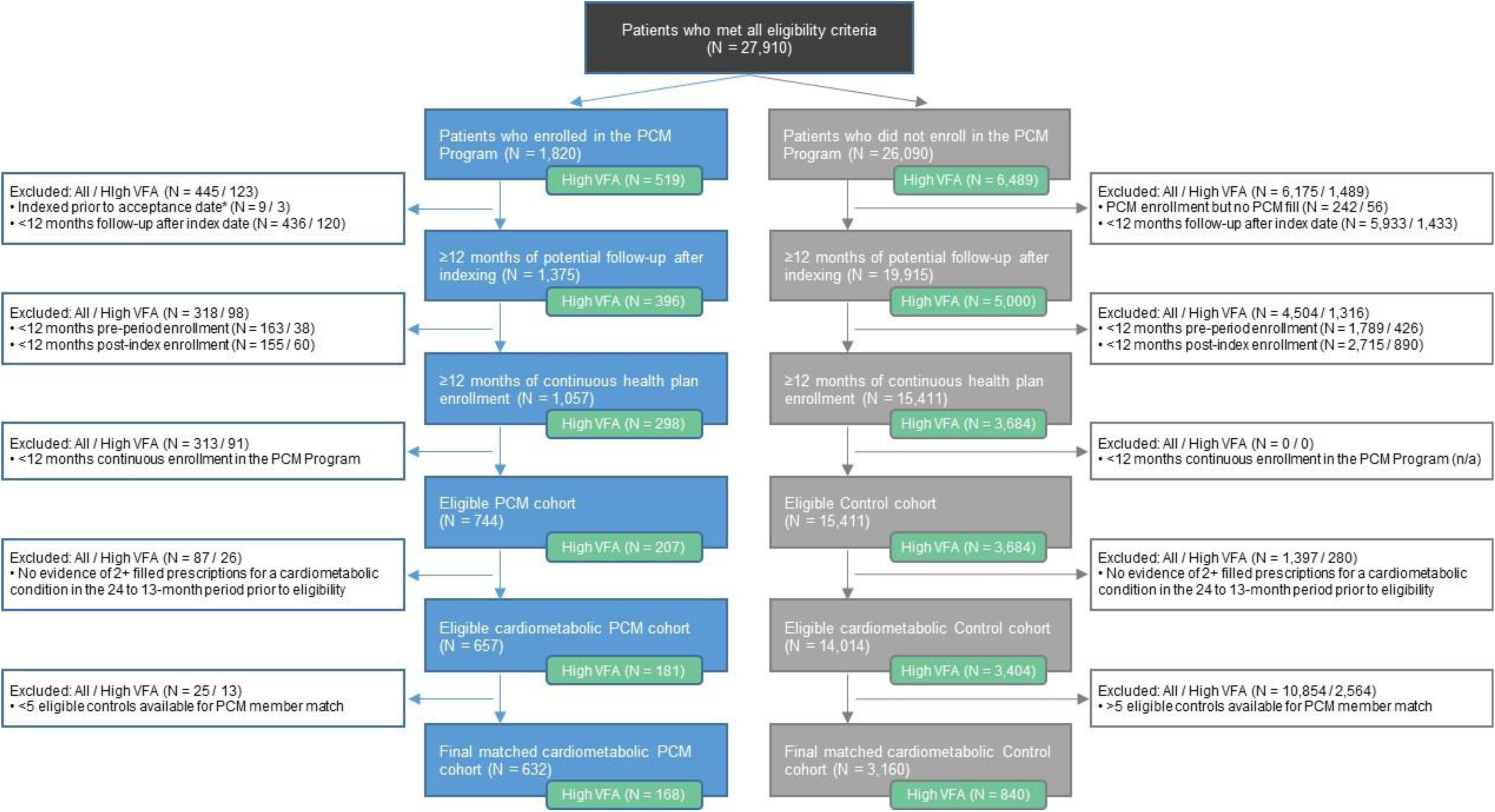
Attrition of patients in the study cohorts. *Indexed prior to acceptance date are those patients who fill a script in the PCM program prior to receiving outreach (e.g., a spouse of a patient was offered the service).

**Table 1.** Baseline characteristics of pharmacy care management (PCM) and control patients with cardiometabolic disease after matching.

| Baseline Characteristics* | PCM<br>(n = 632) | Control<br>(n = 3,160) | SMD |
| --- | --- | --- | --- |
| **Age, Mean (SD), y** | 69.2 (10.1) | 69.5 (10.6) | 0.0264 |
| **Male, No. (%)** | 249 (39.4%) | 1,267 (40.1%) | 0.0142 |
| **Low-income Subsidy Status, No. (%)** | 259 (41.0%) | 1,302 (41.2%) | 0.0045 |
| **Charlson Comorbidity Index Score, Mean (SD)** | 3.6 (2.5) | 3.6 (2.5) | 0.0195 |
| **Comorbid Conditions, No. (%)** |  |  |  |
| Chronic kidney disease | 117 (18.5%) | 655 (20.7%) | 0.0558 |
| Congestive heart failure | 101 (16.0%) | 517 (16.3%) | 0.0103 |
| Coronary heart disease | 214 (33.9%) | 1,092 (34.6%) | 0.0147 |
| Diabetes mellitus (type 2) | 445 (70.4%) | 2,306 (73.0%) | 0.0569 |
| Hypertension | 552 (87.3%) | 2,766 (87.5%) | 0.0057 |
| **Baseline Medication Use** |  |  |  |
| 90-day supply of maintenance medications, no. (%) | 619 (97.9%) | 3,087 (97.7%) | 0.0173 |
| Total maintenance medications, mean (SD) | 10.9 (2.9) | 10.8 (2.8) | 0.0180 |
| Medication classes, no. (%) |  |  |  |
| Anticoagulants | 146 (23.1%) | 782 (24.7%) | 0.0386 |
| Antidepressants | 403 (63.8%) | 1,792 (56.7%) | 0.1446 |
| Anti gout medications | 50 (7.9%) | 260 (8.2%) | 0.0116 |
| Antiplatelets | 129 (20.4%) | 566 (17.9%) | 0.0636 |
| Antipsychotics | 73 (11.6%) | 358 (11.3%) | 0.0070 |
| Beta blockers | 394 (62.3%) | 2,011 (63.6%) | 0.0269 |
| Calcium channel blockers | 268 (42.4%) | 1,298 (41.1%) | 0.0270 |
| Cholesterol lowering agents | 531 (84.0%) | 2,575 (81.5%) | 0.0671 |
| Hypothyroid medications | 202 (32.0%) | 962 (30.4%) | 0.0328 |
| Oral hypoglycemics | 306 (48.4%) | 1,529 (48.4%) | 0.0006 |
| Renin-angiotensin-system-acting agents | 448 (70.9%) | 2,290 (72.5%) | 0.0351 |
| Respiratory medications | 190 (30.1%) | 983 (31.1%) | 0.0227 |
| **Baseline Resource Use** |  |  |  |
| Emergency room visits, mean per 1,000 patients | 77.9 | 72.5 | 0.0392 |
| Inpatient stays, mean per 1,000 patients | 40.2 | 42.0 | 0.0220 |
| Total inpatient days, mean (SD) | 4.1 (11.9) | 4.7 (14.4) | 0.0499 |
| **High VFA Score, No. (%)** | 168 (26.6%) | 840 (26.6%) | 0.0000 |
| **Completed CMR, No. (%)** | 206 (32.6%) | 1,030 (32.6%) | 0.0000 |

PCM = pharmacy care management, CMR = comprehensive medical review; VFA = Value of Future Adherence, SMD = standardized mean difference, SD = standard deviation.
\*Baseline characteristics were assessed during the 12-month period before the index date.

The impact of the PCM program on healthcare resource utilization and medication adherence is shown in **Table 2**. Use of PCM was associated with statistically significant absolute increases in the proportion of patients fully adherent to all classes of cardiometabolic medications, ranging from 4.7% (P = 0.028) for anticoagulants to 17.0% (P < 0.001) for beta-blockers. Improvements in adherence from PCM were particularly large for patients with high baseline VFA scores, ranging from 6.1% (P = 0.032) for anticoagulants to 26.4% (P < 0.001) for beta blockers (**Table e3**).

**Table 2.**
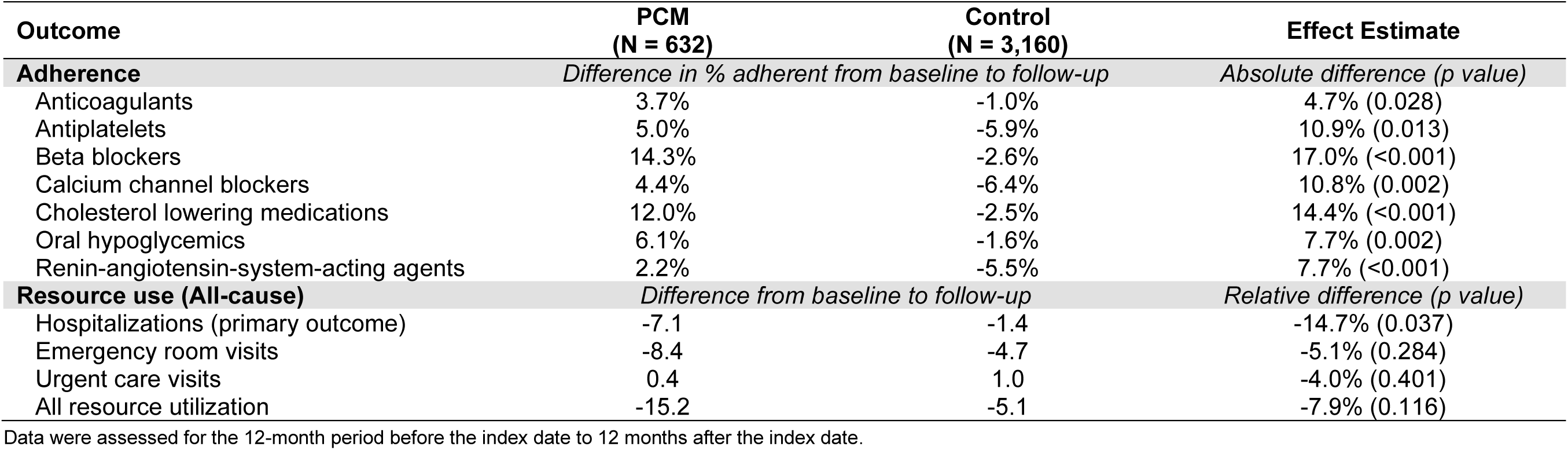
12-month differences in outcomes between PCM and matched controls with cardiometabolic conditions.

| Outcome | PCM<br>(N = 632) | Control<br>(N = 3,160) | Effect Estimate |
| --- | --- | --- | --- |
| **Adherence** | *Difference in % adherent from baseline to follow-up* |  | *Absolute difference (p value)* |
| Anticoagulants | 3.7% | -1.0% | 4.7% (0.028) |
| Antiplatelets | 5.0% | -5.9% | 10.9% (0.013) |
| Beta blockers | 14.3% | -2.6% | 17.0% (<0.001) |
| Calcium channel blockers | 4.4% | -6.4% | 10.8% (0.002) |
| Cholesterol lowering medications | 12.0% | -2.5% | 14.4% (<0.001) |
| Oral hypoglycemics | 6.1% | -1.6% | 7.7% (0.002) |
| Renin-angiotensin-system-acting agents | 2.2% | -5.5% | 7.7% (<0.001) |
| **Resource use (All-cause)** | *Difference from baseline to follow-up* |  | *Relative difference (p value)* |
| Hospitalizations (primary outcome) | -7.1 | -1.4 | -14.7% (0.037) |
| Emergency room visits | -8.4 | -4.7 | -5.1% (0.284) |
| Urgent care visits | 0.4 | 1.0 | -4.0% (0.401) |
| All resource utilization | -15.2 | -5.1 | -7.9% (0.116) |

Data were assessed for the 12-month period before the index date to 12 months after the index date.

The rate of all-cause hospitalization during follow-up was reduced from baseline by 7.1% for PCM patients and decreased by 1.4% for controls. In the difference-in-difference analysis, PCM program participation was associated with a 15% reduction in hospitalizations per 1,000 patient months (*P* = 0.037) (**Table 2**). Reductions in hospitalization rates were particularly large for patients with high baseline VFA scores (PCM -37.2% versus control -18.6%; difference-in-difference -32.0%; P < 0.003) (**Table e3**). Rates of emergency room, urgent care, and the composite of hospitalization, emergency room, and urgent care visits were lower among PCM patients than controls, although these differences were not statistically significant (**Table 2**).

The impact of the PCM program on hospitalization stratified by the reason for admission is shown in **Figure 2**. PCM program enrollees experienced significant reductions in cardiometabolic admissions (difference-in-difference -33.7%; P = 0.025) and non-significant reductions in non-cardiometabolic admissions (difference-in-difference -8.4%; P = 0.201). Although all patients appear to have benefited from the PCM program, reductions in cardiometabolic admissions were largest for individuals who were non-adherent at baseline (**Figure 3**). Specifically, patients who were non-adherent to cardiometabolic medications at baseline had a statistically significant 39.1% (P = 0.003) reduction in cardiometabolic hospitalizations compared to a smaller and non-significant 24.7% reduction (P = 0.286) among those who were adherent at baseline.

**Figure 2.**
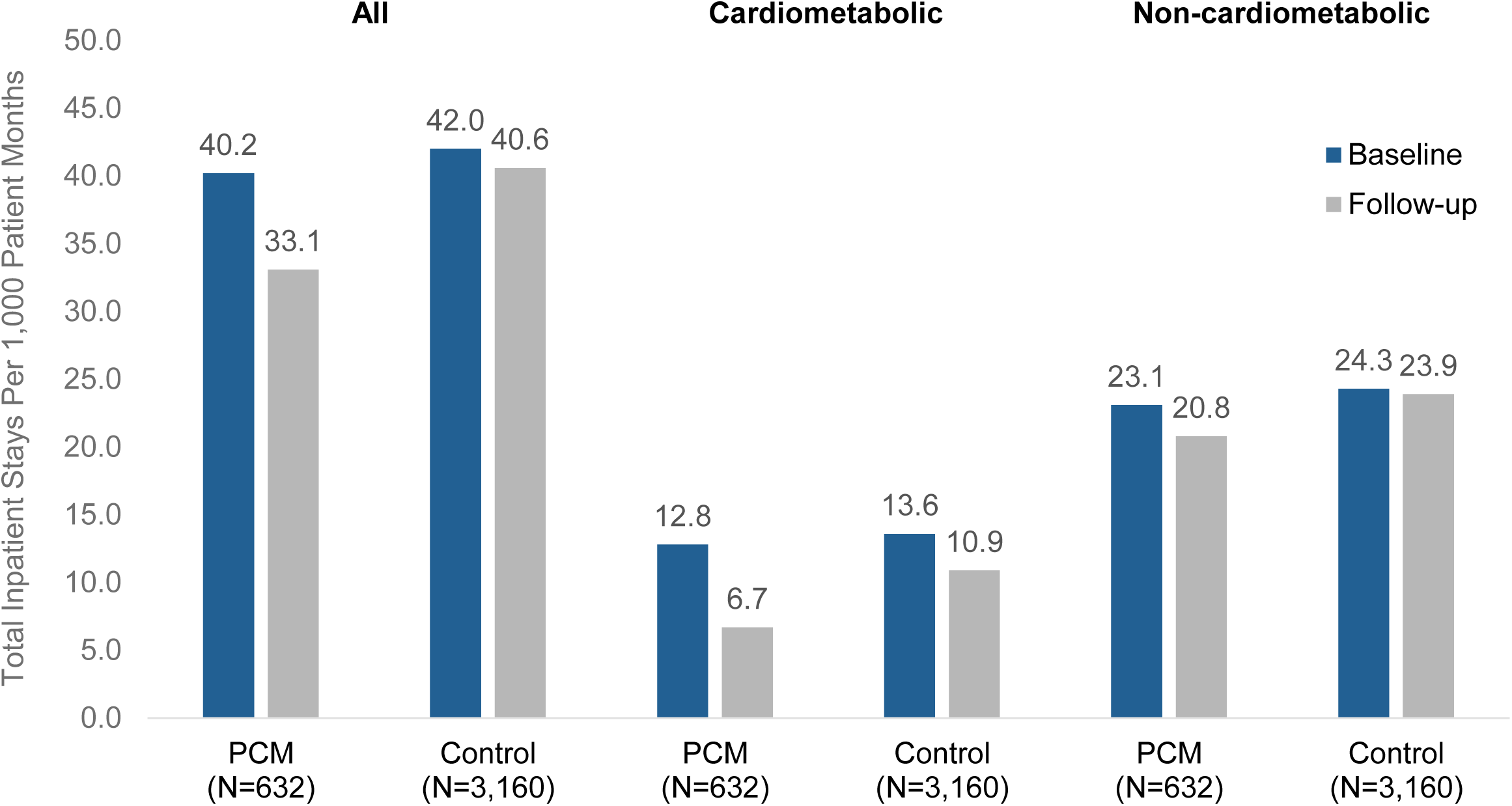
Impact of the pharmacy care management (PCM) program on cardiometabolic and non-cardiometabolic hospitalizations before and after index.

**Figure 3.**
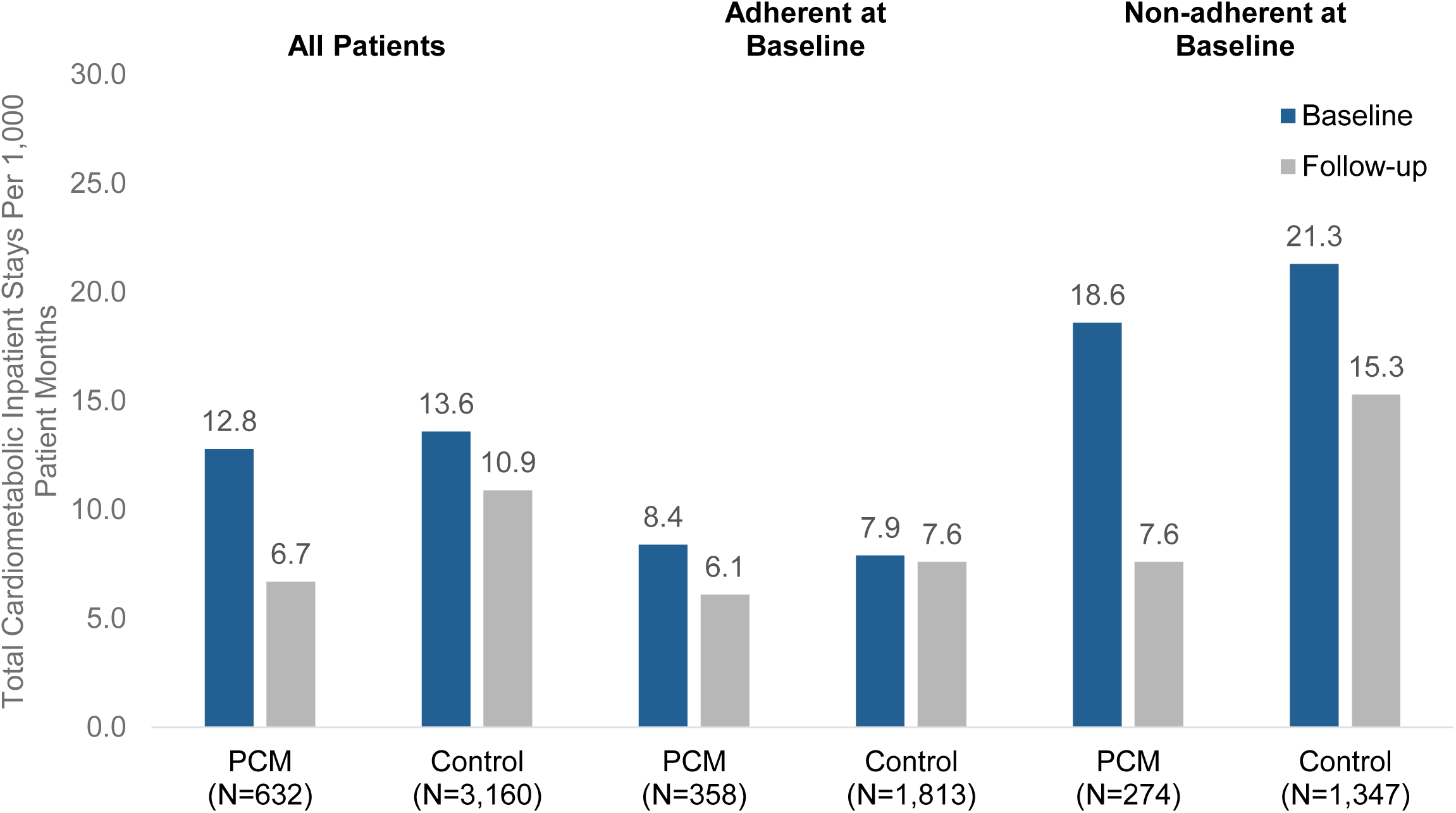
Rate of cardiometabolic hospitalizations between pharmacy care management (PCM) and control patients before and after index, stratified by baseline cardiometabolic medication adherence.

## DISCUSSION

In this propensity-matched cohort study of Medicare beneficiaries with cardiometabolic conditions, we observed that a pharmacy care management (PCM) program significantly increased medication adherence and reduced hospitalizations, predominantly those resulting from cardiometabolic causes. The observed benefits were greatest among enrollees who were non-adherent prior to enrolling in the PCM program.

This PCM program combined multiple evidence-based interventions including medication reconciliation,^25,26^ refill synchronization,^27^ adherence packaging,^28^ monthly medication reviews with member education,^29^ and coordination among prescribing health care providers.^30^ Prior evaluations of PCM programs have demonstrated benefits for a broad range of medically complex patients with high medication burden^12,28^ yet none, to our knowledge have specifically assessed the impact of an ongoing PCM program for patients with cardiometabolic conditions. Given the significant burden of cardiometabolic conditions on the healthcare system ^31,32^ and persistently high rates of non-adherence to effective medications ^33^, this evaluation fills an important gap in the evidence supporting investment in comprehensive PCM programs.

In this context, the clinically meaningful improvements in adherence and economically relevant reductions in hospitalization that we observed have implications to professional societies, health delivery organizations and health insurers, including the Medicare Stars program,^6,34^ which have all devoted substantial resources to improving outcomes for individuals with cardiometabolic disease. Of note, our PCM program was not done in isolation; our results were incremental to the effects of ongoing care management efforts employed by UPMC Medicare Advantage plans to improve quality ratings and lower costs of care, including comprehensive medication reviews, medication adherence campaigns, clinical programs for chronic conditions, accountable care incentives for providers, and case management for medically complex patients. Against this backdrop, these results suggest a comprehensive and integrated pharmacy service tailored to the needs of medically complex and vulnerable individuals with cardiometabolic disease created additional value.

In addition to providing evidence that PCM programs improve adherence and reduce hospitalizations for patients with cardiometabolic disease, our results help explain the mechanism by which PCM programs work. Specifically, the reductions in all-cause hospitalization observed in this cohort were driven primarily by the prevention of cardiometabolic admission and these, in turn, were reduced the most in those who were non-adherent at baseline. Said another way, improvements in medication adherence among those who were non-adherent at baseline led to reductions in cardiometabolic hospitalizations, which was the major contributor to the program’s impact on all-cause hospitalization.

Although less impactful, the program also was effective for patients who were adherent at baseline, though the impact was smaller compared to those who were non-adherent. This may reflect program benefits unrelated to adherence improvements, such as closing care gaps, helping prescribers select more effective regimens, or reducing harmful drug interactions. Conversely, consistent with quality measures used by the Medicare Star Ratings,^35^ we defined patients as being adherent if they had medication available to them 80% of the time. However, for some cardiometabolic medications, such as oral anticoagulants, that require consistent dosing to achieve therapeutic effect, there could be incremental benefits on cardiometabolic outcomes from improved adherence even above this threshold.

It also is worth noting that the PCM program appeared to have a small impact on non-cardiometabolic hospitalization. While the patients in our cohort all had cardiometabolic disease, they were medically complex, had numerous other comorbidities, used many other medications and, in many cases, were socially vulnerable. Since PCM programs take a comprehensive approach to patient management rather than focusing on specific diseases, it is not surprising that patients benefit across a range of conditions by improving treatment regimens, increasing adherence, and deprescribing high-risk medications.

Our study is not without limitations. Our program was deployed in a Medicare Advantage plan. While more than half of Medicare beneficiaries are enrolled in Advantage plans, and this proportion is anticipated to continue to grow,^36^ our findings may not apply to settings with less medically complex or younger patients. In addition, PCM and control patients were not randomly assigned, and the opt-in nature of the program (although required by Medicare regulations) could have led to selection bias. It is reassuring that differences in observed baseline characteristics known to affect resource utilization and adherence were well balanced after direct matching and propensity matching. Although our matched approach with a difference-in-difference adjustment is a robust analytic approach, the potential for unmeasured confounding between the groups remains. The evaluation assessed PCM program efficacy in patients continuously exposed for 12 months, as it takes time for interventions to impact health services utilization, and this duration aligns with similar studies. However, our study does not determine the duration of exposure required for clinical benefit, and results may differ for those exposed for shorter periods.

In conclusion, compared with closely matched controls, we observed significant improvements in medication adherence and reductions in hospitalizations for medically complex and vulnerable Medicare Advantage beneficiaries with cardiometabolic disease, in particular among those with high predicted benefits from improved medication adherence assessed using the VFA score. These benefits were attributable primarily to reductions in admissions for cardiometabolic causes and were especially pronounced among those non-adherent at baseline. Our results provide insights into how PCM programs achieve their benefits and underscore the value of targeting the PCM program to those who are most likely to benefit.

## DISCLOSURES

Shawn Hallinan, Loren Lidsky, Josh Benner, Stephen Jones, Elise Smith, and Niteesh Choudhry receive compensation from RxAnte, Inc. that includes stock options. Chester B. Good has nothing to disclose. Chronis Manolis is an uncompensated member of the RxAnte Board of Directors.

UPMC Enterprises, a division of UPMC, maintains an ownership interest in RxAnte, Inc., which is the owner of RxAnte Pharmacy Services, LLC, (doing business as Mosaic Pharmacy Service), the pharmacy that provided PCM services in this study. UPMC Health Plan has participated in this study by supplying data to RxAnte, Inc. for analysis.

## Data Availability

Supporting data are not available.

## APPENDIX

**Table e1.** Cardiometabolic medications used for cohort entry.

|  |
| --- |
| Anticoagulants |
| Anti gout medications |
| Antiplatelets |
| Beta blockers |
| Calcium channel blockers |
| Cholesterol lowering agents |
| Oral hypoglycemics |
| Renin-angiotensin-system-acting agents |

**Table e2.** Baseline characteristics of pharmacy care management (PCM) and control patients with cardiometabolic disease before matching.

| **Baseline Characteristics\*** | **PCM**<br>**(n = 657)** | **Control**<br>**(n = 14,014)** | **SMD** |
| --- | --- | --- | --- |
| **Age, Mean (SD), y** | 68.9 (10.2) | 72.1 (10.3) | 0.3103 |
| **Male, No. (%)** | 263 (40.0%) | 5,985 (42.7%) | 0.0544 |
| **Low-income Subsidy Status, No. (%)** | 275 (41.9%) | 4,711 (33.6%) | 0.1706 |
| **Charlson Comorbidity Index Score, Mean (SD)** | 3.6 (2.5) | 3.7 (2.6) | 0.0284 |
| **Diagnostic Conditions, No. (%)** |  |  |  |
| Chronic kidney disease | 129 (19.6%) | 3,154 (22.5%) | 0.0705 |
| Congestive heart failure | 110 (16.7%) | 2,470 (17.6%) | 0.0234 |
| Coronary heart disease | 226 (34.4%) | 5,029 (35.9%) | 0.0311 |
| Diabetes mellitus (type 2) | 463 (70.5%) | 10,266 (73.3%) | 0.0619 |
| Hypertension | 576 (87.7%) | 12,244 (87.4%) | 0.0091 |
| **Baseline Medication Use** |  |  |  |
| 90-day supply of maintenance medications, no. (%) | 642 (97.7%) | 12,775 (91.2%) | 0.2891 |
| Total maintenance medications, mean (SD) | 11.0 (3.1) | 10.3 (2.7) | 0.2487 |
| Medication classes, no. (%) |  |  |  |
| Anticoagulants | 158 (24.1%) | 3,710 (26.5%) | 0.0558 |
| Antidepressants | 420 (63.9%) | 7,460 (53.2%) | 0.2184 |
| Anti gout medications | 54 (8.2%) | 1,254 (8.9%) | 0.0260 |
| Antiplatelets | 132 (20.1%) | 2,533 (18.1%) | 0.0513 |
| Antipsychotics | 78 (11.9%) | 1,378 (9.8%) | 0.0656 |
| Beta blockers | 412 (62.7%) | 8,942 (63.8%) | 0.0228 |
| Calcium channel blockers | 277 (42.2%) | 5,885 (42.0%) | 0.0034 |
| Cholesterol lowering agents | 547 (83.3%) | 11,395 (81.3%) | 0.0510 |
| Hypothyroid medications | 208 (31.7%) | 4,097 (29.2%) | 0.0527 |
| Oral hypoglycemics | 316 (48.1%) | 6,236 (44.5%) | 0.0722 |
| Renin-angiotensin-system-acting agents | 462 (70.3%) | 10,013 (71.4%) | 0.0249 |
| Respiratory medications | 198 (30.1%) | 4,004 (28.6%) | 0.0344 |
| **Baseline Resource Use** |  |  |  |
| Emergency room visits, mean per 1,000 patients | 80.8 | 66.0 | 0.1038 |
| Inpatient stays, mean per 1,000 patients | 42.9 | 45.7 | 0.0333 |
| Total inpatient days, mean (SD) | 4.4 (12.1) | 5.6 (16.0) | 0.0827 |
| **High VFA Score, No. (%)** | 181 (27.5%) | 3,404 (24.3%) | 0.0744 |
| **Completed CMR, No. (%)** | 222 (33.8%) | 3,859 (27.5%) | 0.1359 |

PCM = pharmacy care management, CMR = comprehensive medical review; VFA = Value of Future Adherence, SMD = standardized mean difference, SD = standard deviation, CMR = comprehensive medical review. \*Baseline characteristics were assessed during the 12-month period before the index date.

**Table e3.** 12-month differences in outcomes between High Value of Future Adherence (VFA) PCM and matched controls with cardiometabolic conditions.

| Outcome | High VFA PCM<br>(N = 168) | Control<br>(N = 840) | Effect Estimate |
| --- | --- | --- | --- |
| **Adherence** | *Difference in % adherent from baseline to follow-up* |  | *Absolute difference (p value)* |
| Anticoagulants | 8.9% | 2.7% | 6.1% (0.032) |
| Antiplatelets | 5.0% | -7.2% | 12.2% (0.045) |
| Beta blockers | 23.6% | -2.8% | 26.4% (<0.001) |
| Calcium channel blockers | 1.3% | -6.7% | 8.0% (0.130) |
| Cholesterol lowering medications | 14.8% | 1.4% | 13.4% (<0.001) |
| Oral hypoglycemics | 11.3% | 0.3% | 11.0% (0.016) |
| Renin-angiotensin-system-acting agents | 6.1% | -5.7% | 11.7% (0.006) |
| **Resource use (All-cause)** | *Difference from baseline to follow-up* |  | *Relative difference (p value)* |
| Hospitalizations (primary outcome) | -37.2 | -18.6 | -32.0% (0.003) |
| Emergency room visits | -17.9 | -1.9 | -15.9% (0.149) |
| Urgent care visits | -1.5 | 0.6 | -14.4% (0.311) |
| All resource utilization | -56.5 | -19.8 | -21.2% (0.020) |

Data were assessed for the 12-month period before the index date to 12 months after the index date.

## Notes

### Funding Statement

This study did not receive any funding.

### Author Declarations

This study was a program evaluation by UPMC Health Plan and thus deemed exempt by the UPMC Institutional Review Board.

